# Open and laparoscopic approaches are associated with comparable 90-day morbidity and mortality following ERAS protocol

**DOI:** 10.1101/2020.04.08.20057521

**Authors:** Bhavin B. Vasavada, Hardik Patel

## Abstract

**Introduction:** The aim of this study is to compare 90-day mortality and morbidity between open and laparoscopic surgeries performed in one centre since the introduction of ERAS protocols.

**Material and Methods:** All gastrointestinal surgeries performed between April 2016 and March 2019 at our institution after the introduction of ERAS protocols have been analysed for morbidity and mortality. The analysis was performed in a retrospective manner using data from our prospectively maintained database.

**Results:** We performed 245 gastrointestinal and hepatobiliary surgeries between April 2016 and March 2019. The mean age of patients was 50.96 years. 135 were open surgeries and 110 were laparoscopic surgeries. The mean ASA score was 2.4, the mean operative time was 111 minutes and the mean CDC grade of surgery was 2.56. 40 were emergency surgeries and 205 were elective surgeries. Overall the 90-day mortality rate was 8.5% and the morbidity rate was around 9.79%. On univariate analysis morbidity was associated with a higher CDC grade of surgeries, a higher ASA grade, longer operating time, the use of more blood products, a longer hospital stay and open surgeries. HPB surgeries and luminal surgeries (non hpb gastrointestinal surgeries) were associated with 90 day post operative morbidity. On multivariate analysis no factors independently predicted morbidity. On univariate analysis 90-day mortality was predicted by the grade of surgeries, a higher ASA grade, longer operative time, the use of more blood products, open surgeries and emergency surgeries. However on multivariate analysis only the use of more blood products was independently associated with mortality

**Conclusion:** The 90-day mortality and morbidity rates between open and laparoscopic surgeries after the introduction of ERAS protocol were similar.

## Introduction

Early recovery after surgery (ERAS) protocol is becoming the gold standard in perioperative care with excellent results in colorectal, gastric and HPB surgeries.^1^

ERAS is an evidence-based perioperative protocol which has shown significant improvements in perioperative outcomes.^2^ Despite this overwhelming evidence the implementation of these protocols has been very slow and lacks widespread approval.^3^

ERAS was initially developed for colorectal surgeries ^4^, although it is being tested in all other fields.^4^ Unfortunately, it is difficult to have a common ERAS protocol across all subspecialties of GI surgery.

In this paper we evaluate perioperative outcomes since the introduction of ERAS protocols, which includes upper gastrointestinal, HPB and colorectal surgery performed using a laparoscopic or open approach.

Laparoscopic gastrointestinal surgeries have significantly reduced perioperative morbidity and mortalities.^5^ However, many studies reporting these outcomes were conducted before the ERAS era. Very few studies have compared laparoscopic vs open gastrointestinal surgeries since the introduction of ERAS protocols.

## Materials and Methods

Since the introduction of ERAS protocols, data for morbidity and mortality from all gastrointestinal and HPB surgeries performed between April 2016 and March 2019 in our institution have been collected prospectively. Morbidity was defined as any grade of complications according to the Clavien-Dindo classification.^6^ We also evaluated the factors responsible for morbidity and mortalities as well as studies regarding whether there is any difference in 90-day morbidity and mortality between the open or laparoscopic group following ERAS protocol. We perform laparoscopic surgery, unless there is a need for conversion, in all benign and elective cases, while we perform open surgeries in the case of malignant conditions (as we have doubts concerning the oncological outcomes of laparoscopic surgeries in the light of recently published trials) and emergency surgeries.^7^ Ethical approval for our clinical study was obtained from shalby hospitals review board (COA number SBI-5243).

## ERAS protocols

We follow the perioperative guidelines for ERAS protocols described for colorectal surgeries in all our gastrointestinal and HPB surgeries.^8^ Our ERAS protocols are explained in Table 1.

**Table 1.** The ERAS protocol as used in the authors’ centre.

|  |  |
| --- | --- |
| 1. Preoperative course | Nill by mouth not more than 4 hours |
|  | Single dose antibiotics at the time of induction |
| Intra-operative and post-operative Course | No ryle's tube post operatively |
|  | No post operative antibiotics except ongoing sepsis |
|  | Early ambulation within 6 hours |
|  | Liquid orally within 6 hours, soft diet on post-operative day 1 and normal diet according to patient's wish from post-operative day 2 |
|  | No intraoperative drains |
|  | Day 1 foley's catheter removal. |

## Statistical analysis

Statistical analysis was performed using SPSS version 23. A p value less than 0.05 was considered statistically significant. The categorial values were evaluated using the chi square test, while the numerical values were evaluated using the Mann-Whitney U test. The multivariate analysis was performed using the logistic regression method. A Cox multivariate regression analysis was performed to evaluate the factors effecting 90-day overall and morbidity-free survivals. All the factors with a p value of less than 0.05 were included in the multivariate analysis.

## Results

We performed 245 gastrointestinal and hepatobiliary surgeries between April 2016 and March 2019. The mean age of patients was 51 years. 144 patients were male and 101 were female. 135 were open surgeries and 110 were laparoscopic surgeries. 11 were upper gastrointestinal surgeries (stomach and oesophagus), 27 were small intestinal surgeries, 143 were HPB surgeries, 38 colorectal and 26 hernia surgeries. The mean ASA score was 2.4, the mean operative time was 111 minutes, and the mean CDC grade of surgery was 2.56. 40 were emergency surgeries and 205 were elective surgeries.

The overall 90-day mortality rate was 8.5% and the overall morbidity rate was 9.79%. The 90-day mortality and morbidity in elective surgeries was 4.08% and 8.16% respectively.

On univariate analysis 90-day morbidity was significant with a higher CDC grade of surgeries, a higher ASA grade, longer operative time, the use of more blood products and a longer hospital stay. HPB and luminal (stomach, small intestine and large intestine) surgeries were associated with morbidity longer than 90 days(Table 2). On multivariate analysis no factor independently predicted morbidity.

**Table 2.** Univariate and multivariate analysis of morbidity.

| Factors | Morbidity<br>Present<br>(n=24) | Morbidity<br>Absent<br>(n=221) | Univariate<br>analysis (P<br>value) | Multivariate<br>analysis<br>(Pvalue) |
| --- | --- | --- | --- | --- |
| Open surgery vs<br>Laprosopic | Lap:<br>1,Open:2<br>3 | Lap:<br>109,Open:<br>112 | <0.0001 | 0.091 |
| Emergency surgery<br>(N)= 40 | 4 | 36 | 1.0 |  |
| Luminal<br>Surgery(non HPB<br>gastrointestinal<br>surgery) | 16 | 59 | <0.0001 | 0.377 |
| HPB surgery | 6 | 133 | 0.003 | 0.573 |
| Age(mean) | 51.17 | 50.94 | 0.850 |  |
| Hospital stay<br>(mean) | 4.5 | 2.95 | <0.0001 | 0.421 |
| CDC grade of<br>surgery (mean) | 3.04 | 2.51 | <0.0001 | 0.962 |
| Blood product | 1.42 | 0.51 | <0.0001 | 0.981 |
| (mean) |  |  |  |  |
| ASA grade (mean) | 2.83 | 2.35 | <0.0001 | 0.691 |
| Operative time<br>(mean) | 168.7 | 104.7 | <0.0001 | 0.142 |

On univariate analysis 90-day mortality was predicted by the grade of surgeries, a higher ASA grade, longer operative time, the use of more blood products, open surgeries and emergency surgeries (Table 3). However, on multivariate analysis the number of blood products used predicted mortality independently (p= 0.046, odds ratio 1.52,95 percent C.I. 1.008-2.317).

**Table 3:** Univariate and multivariate analysis of 90-day mortality.

| Factors | Mortality<br>(n=21) | No mortality<br>(n=224) | Univariate<br>analysis<br>(P value) | Multivariate<br>analysis (P<br>value) |
| --- | --- | --- | --- | --- |
| Emergency<br>surgery(n=21) | 11 | 10 | <0.0001 | 0.116 |
| Open surgery<br>(135) vs<br>laproscopic<br>surgery (110) | Open surgery<br>21/laparoscopic<br>surgery 0 | 114<br>open surgery/11<br>0 laparoscopic<br>surgery | <0.0001 | 0.996 |
| Luminal (non<br>HPB) surgeries | 11 | 159 | 0.087 |  |
| Morbidity<br>(90days) | 5 | 19 | 0.041 | 0.716 |
| Hospital stay<br>(mean) | 3.42 | 3.08 | 0.173 |  |
| Age (mean) | 51.76 | 50.89 | 0.956 |  |
| Blood products<br>(mean) | 2.24 | 0.44 | <0.0001 | 0.046,odds<br>ratio 1.52(95<br>percent C.I.<br>1.008-2.317) |
| CDC grade of<br>surgery (mean) | 3.29 | 2.5 | <0.0001 | 0.818 |
| Operative time<br>(mean) | 177.14 | 104.79 | <0.0001 | 0.263 |
| ASA (Mean) | 3.1 | 2.33 | <0.0001 | 0.656 |

We also performed a univariate and multivariate analysis of different factors between the open and laparoscopic groups.

On univariate analysis open surgeries were associated with increased morbidity and mortality, although on multivariate analysis open surgery did not independently predict morbidity. However, the multivariate analysis showed that in the case of open surgeries operative time is significantly increased.(Table 4).

**Table 4:** multivariate analysis between open and laparoscopic surgeries

| Multivariate analysis of open and laproscopic surgeries |  |  |  |  |  |  |  |  |
| --- | --- | --- | --- | --- | --- | --- | --- | --- |
|  | B | S.E. | Wald | df | Sig. | ODDS RATIO | 95% C.I. for EXP(B) |  |
|  |  |  |  |  |  |  | Lower | Upper |
| Step 1<sup>a</sup> |  |  |  |  |  |  |  |  |
| morbidityall(1) | -1.063 | 1.319 | .650 | 1 | .420 | .345 | .026 | 4.584 |
| mortality_A(1) | -21.629 | 6941.449 | .000 | 1 | .998 | .000 | .000 | . |
| LUMINNAL(1) | .737 | .618 | 1.425 | 1 | .233 | 2.090 | .623 | 7.013 |
| EMMERGENCY(1) | 2.244 | 1.118 | 4.030 | 1 | .045 | 9.433 | 1.055 | 84.371 |
| operativetime | -.016 | .005 | 8.739 | 1 | .003 | .984 | .973 | .995 |
| ASA | 1.330 | .624 | 4.546 | 1 | .033 | 3.780 | 1.113 | 12.836 |
| hospiitalstay | 2.232 | .415 | 28.990 | 1 | .000 | 9.318 | 4.135 | 20.996 |
| gradeofsurgery | .941 | .544 | 2.986 | 1 | .084 | 2.562 | .881 | 7.446 |
| Constant | 10.725 | 6941.450 | .000 | 1 | .999 | 45479.336 |  |  |

a. Variable(s) entered on step 1: morbidityall, mortality\_A, LUMINNAL, EMMERGENCY, operativetime, ASA, hospiitalstay, gradeofsurgery.

To confirm our findings, apart from logistic regression we also performed a multivariate Cox regression analysis of 90-day survivals. In this analysis we found that a lower operative time, a shorter hospital stay and a smaller number of transfused blood products independently predicted a 90-day survival.

We did the same for 90-day morbidity-free survival, which showed that no factor independently predicted morbidity-free survival after the Cox regression multivariate analysis.

We also prepared 90-day morbidity-free and overall survival-rate curves for open and laparoscopic surgeries (Figures 1 and 2), which show that the results of the Cox multivariate analysis are not statistically signifiant.

**Figure 1:**
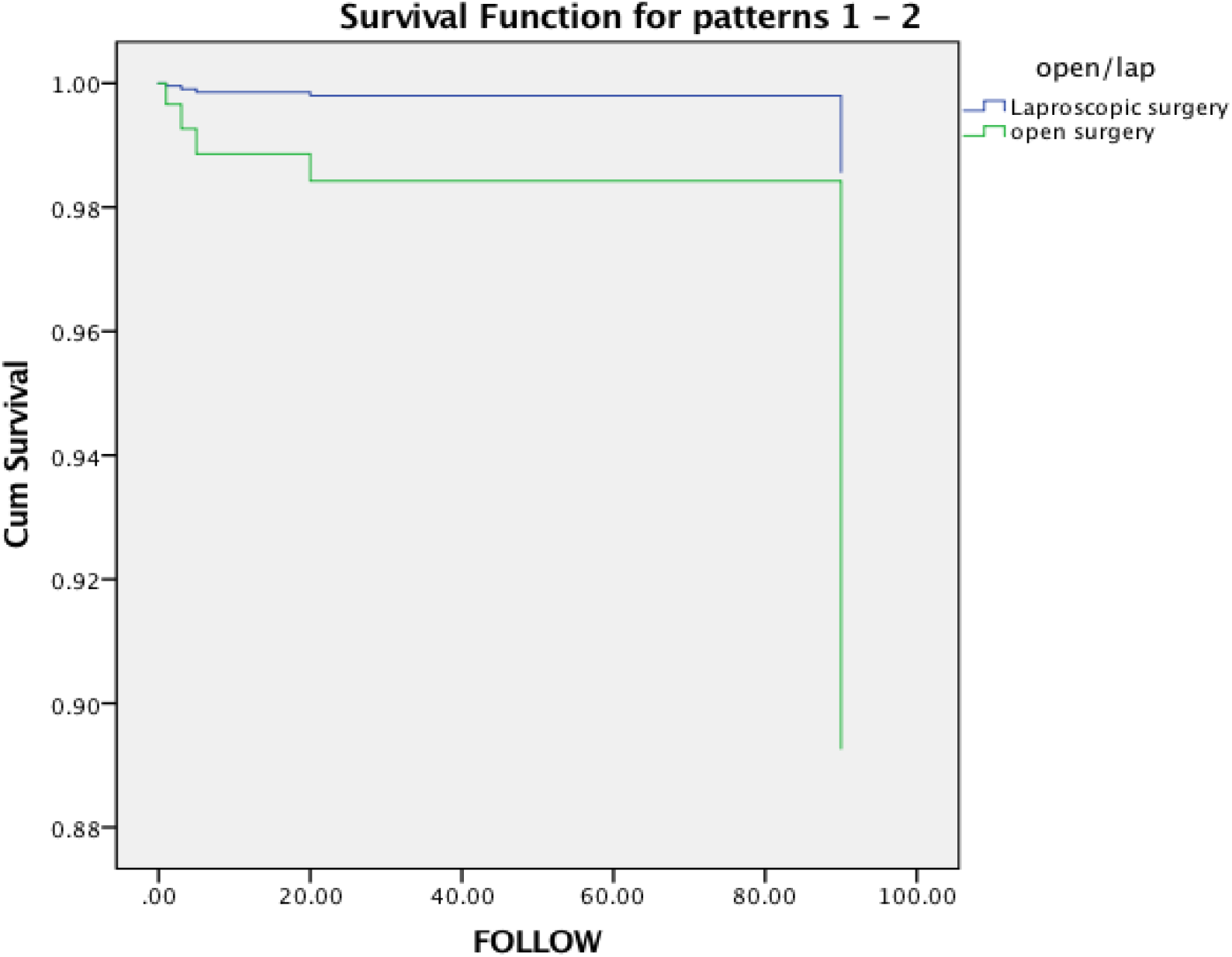
90-day morbidity-free survival between open and laparoscopic surgeries, which did not show statistical significance on multivariate analysis. (p= 0.059, hazard ratio 0.127 95% C.I (0.015-1.079)

**Figure 2:**
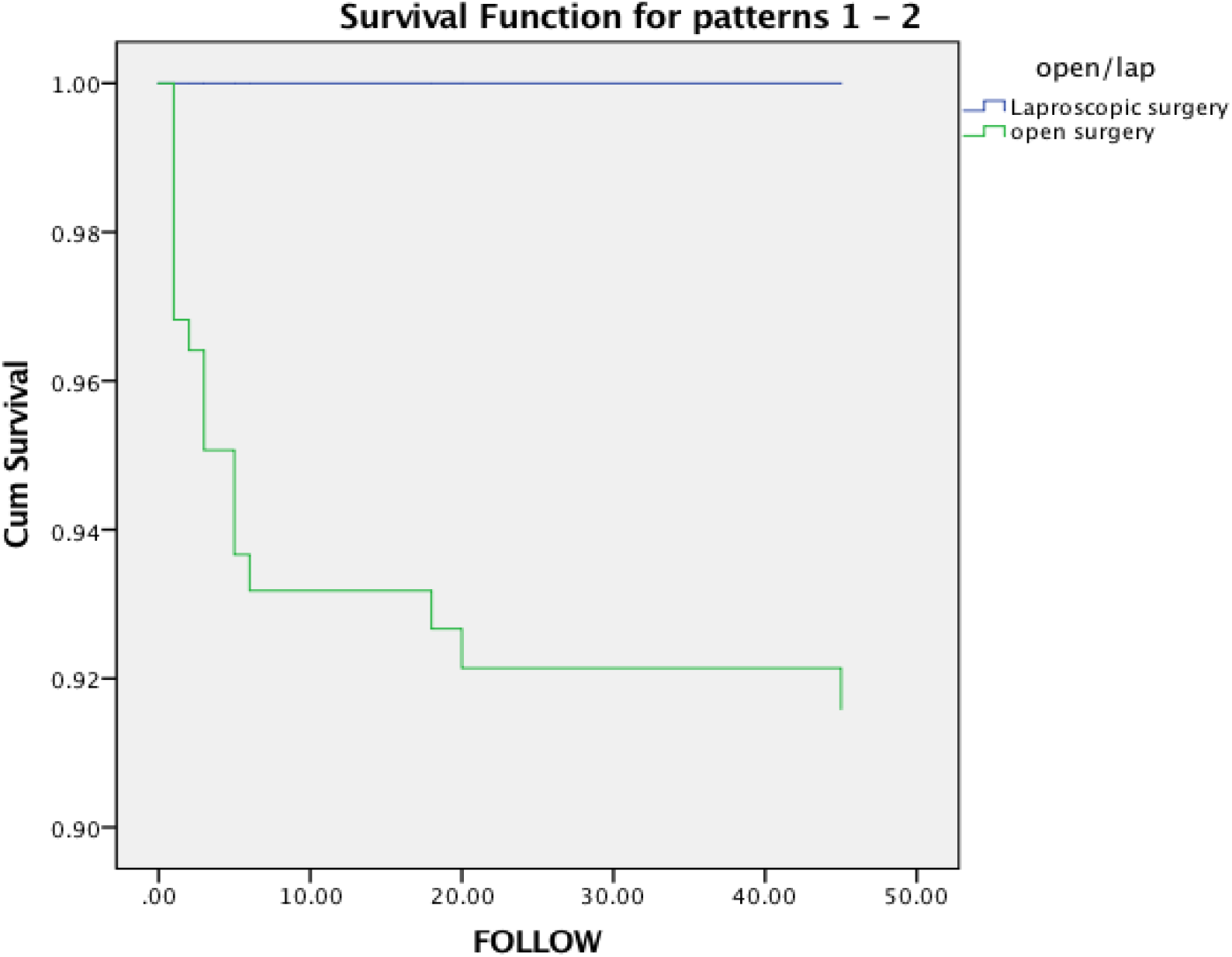
Kaplan meier 90-day survival curve between open and laparoscopic surgery, which did not show a significant difference in the multivariate analysis. (p= 0.920)

## Discussion

Enhanced recovery after surgery, although it was initially described for colorectal surgery, is now becoming standard protocol for all surgeries and it has significantly reduced hospital stays and costs without affecting morbidity and mortality.^1-5^

However, there are very few studies available that study the effect of ERAS protocols on morbidity and mortality in the entire cohort including HPB and Gastrointestinal surgeries.

We follow ERAS protocols for all gastrointestinal and HPB surgeries, as shown in table 1, so we evaluated 90-day morbidity and mortality in our series after implementing ERAS protocols and also studied the various factors affecting 90- day morbidity and mortality.

We also evaluated the difference in morbidity and mortality in the laparoscopic vs open surgery group after implementing the ERAS protocol.

In our series the overall 90-day mortality rate was 8.5% and the overall morbidity rate was around 9.79%. 90-day mortality and morbidity in elective surgery were around 4.08 and 8.16 percent respectively. Mortality is defined as any death within 90 days of the operation and morbidity included Clavien-Dindo grade 3 and grade 4 complications, which is similar to the published data. ^9,10^

In our univariate analysis morbidity was significantly associated with a higher CDC grade of surgeries, a higher ASA grade, a longer operative time, the use of more blood products, a longer hospital stay and open surgeries. HPB surgeries and luminal surgeries (non hpb gastrointestinal surgeries) were associated with higher 90-day morbidity. However, in the multivariate analysis no factors independently predicted morbidity.

In the univariate analysis 90-day mortality was predicted by the grade of surgeries, a higher ASA grade, a longer operative time, greater requirements for blood products, open surgeries and emergency surgeries and in the multivariate analysis only the greater use of blood products was independently associated with higher mortality. These data show that laparoscopic or open surgeries did not predict 90-day morbidity and mortalities independently.

Thus, 90-day morbidity and mortality analysis in our series shows patient and disease-related factors, which is also shown in various published studies. ^11.12.13^ One of the main aims of our study is to evaluate the 90-day morbidity and mortality difference between open and laparoscopic surgeries after implementing the ERAS protocol.

Various published studies show that morbidity after colorectal resection was lower in laparoscopic than in open patients.^14,15^ In the case of cholecystectomy it is now concluded that laparoscopic surgery reduces morbidity. ^16^ However, all these papers were published before the widespread introduction of the ERAS protocol.

In our study we have analysed laparoscopic and open surgery using univariate and multivariate logistic regression [Table 1] analysis. 90-day morbidity and mortality were associated with open surgery in the univariate analysis but multivariate logistic regression showed that open or laparoscopic surgeries did not independently predict 90-day morbidity and mortality after following the ERAS protocol across gastrointestinal and HPB surgeries.

Open surgeries were more complex in terms of longer operative time, the higher number of blood products used, the high CDC grade of surgeries, emergency surgeries and high ASA grades as we perform open surgeries as part of emergency surgeries and oncologic surgeries, which explains their association with morbidity and mortality in univariate analysis. On multivariate analysis open surgeries did not predict 90-day morbidity and mortality independently, which confirms that patient-related factors, rather than the open or laparoscopic approaches to surgeries, predict mortality and morbidity following our ERAS protocol.

To further confirm our findings we performed a 90-day survival analysis on the multivariate cox regression analysis. However, the hospital stay was significantly longer in the open group

Spanjersberg et al ^17^ show that even after implementing ERAS, laparoscopic surgeries were associated with reduced morbidity and a shorter hospital stay, which is contrary to our data, including all kinds of gastrointestinal and HPB surgeries. Zhuang et al^18^ show that the benefit of laparoscopic surgery in optimal ERAS settings is yet to be proved. Damania et al show that the ERAS protocol reduced hospital stays in the case of HPB surgeries, although they did not compare open vs laparoscopic surgeries.

The majority of these data are available for colorectal surgery only. To our knowledge ours is one of the first studies to show that if HPB and gastrointestinal surgeries are considered together, there is no difference in 90- day morbidity and mortality between open and laparoscopic surgeries. The hospital stay was still significantly longer in the case of open surgery.

Our study suffers from the bias inherent in all retrospective studies, and further randomised control trials are needed to confirm our findings. Furthermore, although the primary aim of the study is to evaluate the effect of the ERAS protocol in heterogenous populations, more data for each specific subgroup are needed and we are in the process of evaluating such data.

In conclusion, we have found no difference in 90-day morbidity and mortality between open and laparoscopic surgeries after implementing ERAS protocols, morbidity and mortality being associated with patient and disease-related factors.

## Data Availability

we have all the available data and can produce on demand.

## Abbreviations

ERAS: Enhanced recovery after surgery
HPB: Hepato Pancreato Biliary
GI: Gastrointestinal
CDC: Centers for Disease Control and Prevention
ASA: American Society of Anaesthetists

